# PREDICTORS OF UNMET NEEDS OF FAMILY PLANNING AMONG WOMEN IN THEIR REPRODUCTIVE AGES LIVING WITH HIV IN SUB-SAHARAN AFRICA: ANALYSIS OF DEMOGRAPHIC HEALTH SURVEY

**DOI:** 10.1101/2024.08.29.24312800

**Authors:** Munawar Harun Koray, Mansour Maulid Mshenga, Epafra Luka Mwanja, Sara Sulieman, Damien Punguyire, Augustine Adomah-Afari

## Abstract

**Background:** Unmet needs for family planning (FP) among women of reproductive age living with HIV (WRLWH) increase the risk of mother-to-child transmission (MTCT) of HIV, hindering efforts to control the epidemic. These unmet needs lead to unintended pregnancies among high-risk mothers, resulting in maternal health complications, and risk of MTCT of HIV. This study examines factors predicting unmet FP needs among WRLWH in Sub-Saharan Africa (SSA).

**Method:** This cross-sectional study used secondary data from the Demographic Health Survey (DHS) program, encompassing 14 SSA countries and including 9395 WRLWH. Data analysis, conducted with STATA version 18, involved three steps: computing unmet FP needs using descriptive statistics, assessing associations through Pearson’s chi-square test, and performing multi-level regression analysis to identify predictors of unmet FP needs. Regression models were fitted, considering individual factors, FP exposure, and partner/community factors, with statistical significance set at p-value ≤ 0.05 at a 95% confidence interval. Both random effects and fixed effects were reported.

**Results:** The findings revealed that 15.57% of the WRLWH had unmet FP needs. Older aged women with HIV had lower odds of unmet FP needs, particularly the 45-49 age group (aOR = 0.159, CI = 0.0913-0.277). Higher parity increased the odds, especially for those with four or more children (aOR = 8.081, CI = 5.113-12.77). Exposure to FP through media reduced the odds (aOR = 0.564, CI = 0.468-0.678). Female household heads had higher odds (aOR = 1.306, CI = 1.086-1.571) of unmet need of FP, while rural residents had lower odds (aOR = 0.696, CI = 0.560-0.866) compared to urban residents.

**Conclusion:** The study highlights the necessity for tailored FP programs and targeted media-based education to reduce unmet needs among WLWH, enhance health outcomes, prevent MTCT of HIV, and achieve related SDGs among women in their reproductive age living in SSA.

## BACKGROUND

The Human Immunodeficiency Virus (HIV) is a persistent global health issue affecting millions of people worldwide (1). As of 2022, about 39 million people worldwide were living with HIV, of them 25.6 million were from Africa region. Out of 39 million people living with HIV globally, 37.5 million of them were adults from 15 years of age or older of whom 20.0 million were women (2). The sub Saharan Africa (SSA) occupied about two-thirds of the total number of people living with HIV compared to the rest of the world (3). In the region, women continue to be disproportionately affected and are the most severely affected group (3).

The family planning (FP) for women living in their reproductive age living with HIV (WRLWH) is one of crucial interventions to halting the HIV epidemic (4). Family planning provides benefit of timing and spacing of their pregnancies and prevention of HIV transmission from mother to child (MTCT) for women living with the virus who do not wish to become pregnant (5). Additionally, FP lowers the chance of unintended births among high risk mothers which could result in maternal morbidity, mortality or complications during labor (6).

The unmet need for FP refers to fecund and sexually active women who describe not desiring or delaying having children in the future, and who do not use any form of contraception (7). The relationship between women’s aspirations towards reproduction and their use of contraception is highlighted by the notion of unmet need (8). Women’s capacity to achieve their intended birth spacing and family size can be measured by their unmet FP needs. Additionally, it indicates how well interventions for reproductive health have addressed the need for services (9).

The intersection of HIV and the unmet need for FP has significant implications for the health and well-being of WLWH, as well as for broader public health outcomes (10). Despite the availability of antiretroviral therapy provided at most maternal health centers, the integration of FP and HIV services remains suboptimal, leading to a significant unmet need for contraception (11). This unmet need contributes to higher rates of unintended pregnancies, which complicates the health outcomes for women living with HIV and their children (12, 13).

The unmet need for FP among WRLWH has profound implications for maternal and child health, HIV transmission rates, and overall well-being of infected mothers (13). With the high prevalence of HIV in this region, it is essential to address the unique challenges and barriers faced by WRLWH when it comes to accessing and utilizing FP services (14). Addressing this unmet need is not only a matter of reproductive rights but also a crucial component of HIV prevention and treatment efforts (15). In this context, it is imperative to understand and address the gaps of unmet need among WLWH in SSA (16).

This study therefore aimed to shed light on the factors that has the potential of influencing the unmet needs of FP among a key population of WLWH in SSA. This is crucial for improving public health interventions in the region as it will help to explore potential strategies and interventions to address this pressing issue around SSA (17). The findings will also contribute to ensuring universal access to sexual and reproductive health-care especially FP for WRLWH as enshrined in the Sustainable Development Goal (SDG) 3 (target 3.1, 3.7 and 3.8) and goal 5 (target 5.6) (18) in SSA. Ultimately, this study will contribute to the development of targeted and effective interventions that can empower WRLWH to make informed decisions about their reproductive futures and improve maternal and child health outcomes.

## METHODS

### Study Design and Data Source

This study is a cross-sectional study using secondary data from a nationally representative surveys conducted by the Demographic Health Survey program. The surveys are undertaken in 85 low-and middle-income countries around the world almost every five years (19). About 5000 to 32000. A multistage sampling technique is used to recruit survey respondents. Detailed description of the sampling techniques is described in the DHS website https://www.dhsprogram.com/Methodology/Survey-Types/DHS-Methodology.cfm (19) and literature (20). The data collected includes maternal and child health related indicators. Data for this study was extracted from the individual and anthropometry datasets. Data from the respondents is collected using standardized structured questionnaires and blood samples taken for various tests, including HIV test. The data for this study is freely available in the DHS website https://www.dhsprogram.com/data/available-datasets.cfm (19).

This study population had 115,124 women in their reproductive ages in 14 countries in SSA whose blood samples were taken for HIV testing and were returned with either positive or negative HIV results. The study then included only women in their reproductive ages who test positive of HIV (n = 9395). Table 1 shows both the samples from each country.

**Table 1:**
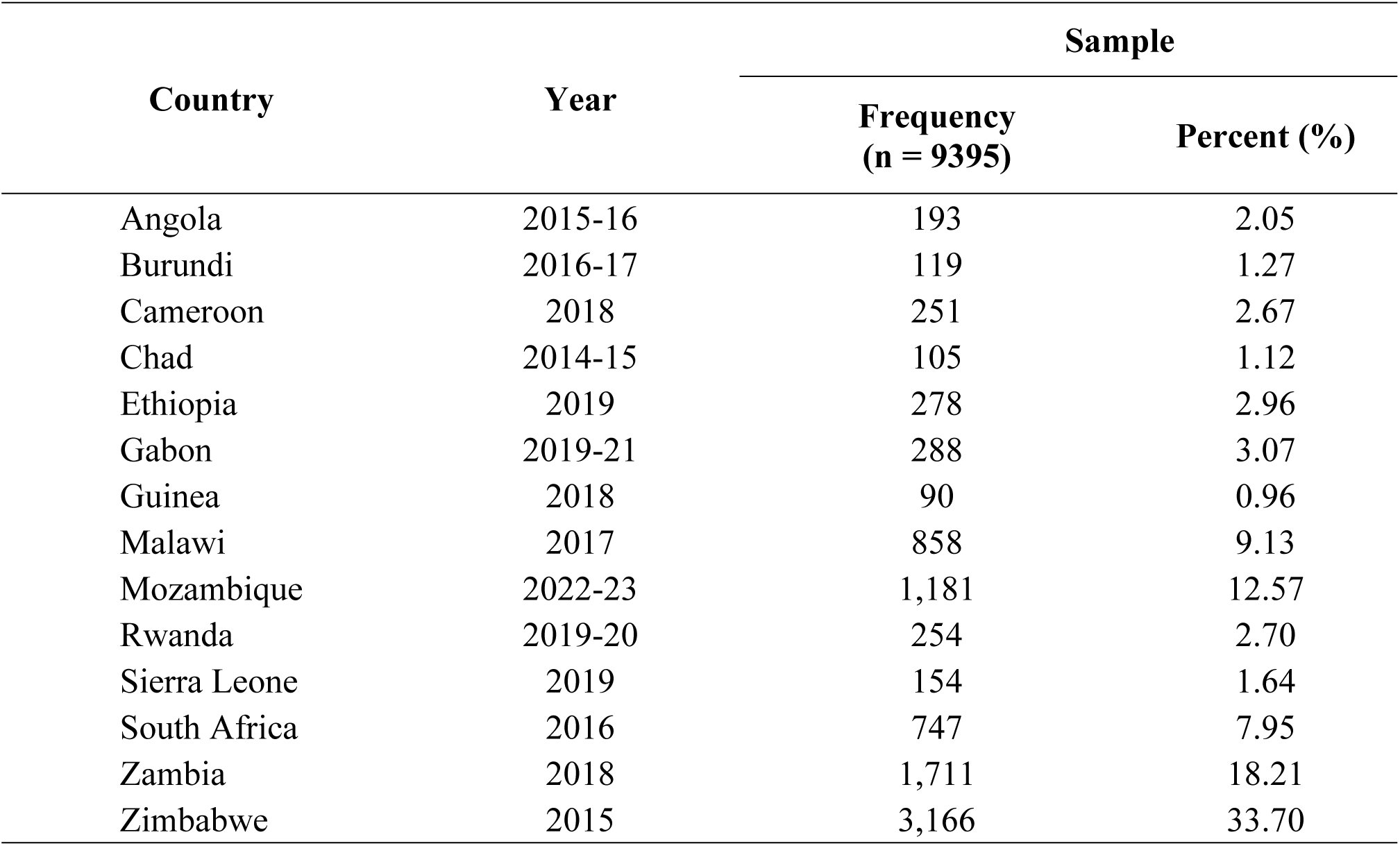
Unweighted and weighted samples from each country.

| Country | Year | Sample |  |
| --- | --- | --- | --- |
|  |  | Frequency<br>(n = 9395) | Percent (%) |
| Angola | 2015-16 | 193 | 2.05 |
| Burundi | 2016-17 | 119 | 1.27 |
| Cameroon | 2018 | 251 | 2.67 |
| Chad | 2014-15 | 105 | 1.12 |
| Ethiopia | 2019 | 278 | 2.96 |
| Gabon | 2019-21 | 288 | 3.07 |
| Guinea | 2018 | 90 | 0.96 |
| Malawi | 2017 | 858 | 9.13 |
| Mozambique | 2022-23 | 1,181 | 12.57 |
| Rwanda | 2019-20 | 254 | 2.70 |
| Sierra Leone | 2019 | 154 | 1.64 |
| South Africa | 2016 | 747 | 7.95 |
| Zambia | 2018 | 1,711 | 18.21 |
| Zimbabwe | 2015 | 3,166 | 33.70 |

### Study Variables

This study is set to investigate the factors associated with unmet needs of FP among WRLWH in SSA. The outcome variable for this study is unmet needs of FP among HIV positive women in their reproductive age. Unmet needs of FP are described using two other measures; 1. Unmet need for spacing, which implies fertile women in childbearing age who would want to space their birth intervals but are currently not using any form of modern contraceptive method, and 2. Unmet needs for limiting birth, which refers to fertile women of childbearing age who wants no additional children but are currently not on any modern contraceptive method.

The explanatory variables were categorized under woman’s factors (age, marital status, religion, parity, educational status, occupation, and wealth index) exposure to FP (exposure to FP through media, knowledge of any contraceptive, knowledge of modern contraceptive, ever used any contraceptive, exposure to FP through radio, exposure to FP through newspaper/magazine) and partner/community factors (partner’s age, husband/partner’s educational level, husband/partner’s occupation, sex of household head, type of place of residence and distance to health facility). These variables were highlighted as significant predictors of unmet needs of FP in earlier studies (21, 22).

### Data Analysis

The data analysis for this study was performed using STATA version 18 macOS, following three-step analytic approach. In the first step, the unmet needs of family planning, including unmet needs of limiting and spacing were computed. Additionally, the association between HIV status and unmet needs of FP were assessed using Pearson’s chi-square test at a p-value ≤ 0.05. In the second stage of the analysis, the prevalence of unmet needs of FP (total) across the independent variables and their association were assessed, again using Pearson’s chi-square test. In the third stage of the analysis, independents variables which had statistically significant association with unmet needs of FP of p < 0.25 were included in a multi-level regression analysis. The multi-level analysis was conducted to compensate for the different levels used in this study (individual factors, exposure to media, partner/community factors). The multilevel regression analysis included mixed and random effects (23).

Four regression models, excluding a null model (model 0) were fitted to identify the predictors of unmet needs of FP among women living with HIV. Model 1 contained individual variables, model 2 had variables on exposure to FP and contraceptive use, the Model 3 had variables on the partner/community factors. All models were set at a p-value ≤ 0.05, at a 95% confidence interval (CI). The last model, model 4, contained all the variables in all the models. Sample weight and SVY command were used to correct for over and under-sampling and the complex survey design and generalizability of the findings respectively.

### Model diagnostics

Variance inflation factor (VIF) was used to assess the presence of multi-collinearity of the model. The presence of the variables “knowledge of any contraceptive method” and “knowledge of modern method” showed collinearity, and therefore were removed. The final model showed presented no evidence of multi-collinearity (mean VIF = 2.03, min = 1.04 and max = 4.22). The STATA commands “mixed” and “fitstat” were used in fitting these models. The variance at the PSU (Primary Sampling Unit) level is fairly consistent across models, suggesting some level of clustering within PSUs.

The models’ comparison was done using and Akaike’s Information Criterion (AIC) tests. The ICCs are low (0.033 to 0.036), indicating minor variability at the primary sampling unit (PSU) level relative to the individual variability. The models improve in their fit from Model 0 to Model 4, as evidenced by decreasing AIC values, suggesting that adding more variables helps in better capturing the factors associated with unmet FP needs among women living with HIV. The likelihood ratio (LR) tests and Wald chi-square tests across the models are highly significant (p < 0.0001), validating the model structures and the inclusion of random effects.

### Ethical Consideration

This study employed the DHS database, which is an international survey conducted over a period of five years. After registering and submitting the topic through their website, ICF International granted permission to access and use the dataset. Detailed information regarding the methodology and ethical considerations can be found on the DHS website. The data used in this study were secondary and publicly accessible, therefore individual consent was not required by the ICF International Institutional Review Board. This study proceeded with the necessary permissions from the DHS Programme, and all data were carefully managed to ensure privacy during the processing and analysis stages of the study.

## RESULTS

It is observed that 17.1% of the entire women included in the study had unmet need of family planning. The HIV status of the women was strongly (p < 0.0001) associated with unmet needs of FP for limiting and spacing of birth and both (total). The study also found that 6.6% of HIV positive women had unmet need of family planning for both limiting and spacing (Table 2).

Figure 1 shows a pooled prevalence of 8.917% unmet need for spacing among WRLWH in SSA, with Angola presenting the highest unmet need for spacing.

Figure 2 presents the pooled prevalence of 5.183% unmet need for limiting among WRLWH in SSA, with Sierra Leone presenting the lowest. For the total unmet need for both spacing and limiting, the pooled prevalence for all 14 countries was 15.570%, with Angola and Burundi recording the highest and lowest total unmet needs for family planning, respectively (Figure 3). There are considerable variations in the prevalence of total unmet needs of family planning (τ^2^ = 23.93, I^2^ = 92.43%, H^2^ = 13.2) among the countries with an overall significant effect (z = 10.80, p < 0.0001).

**Figure 1.**
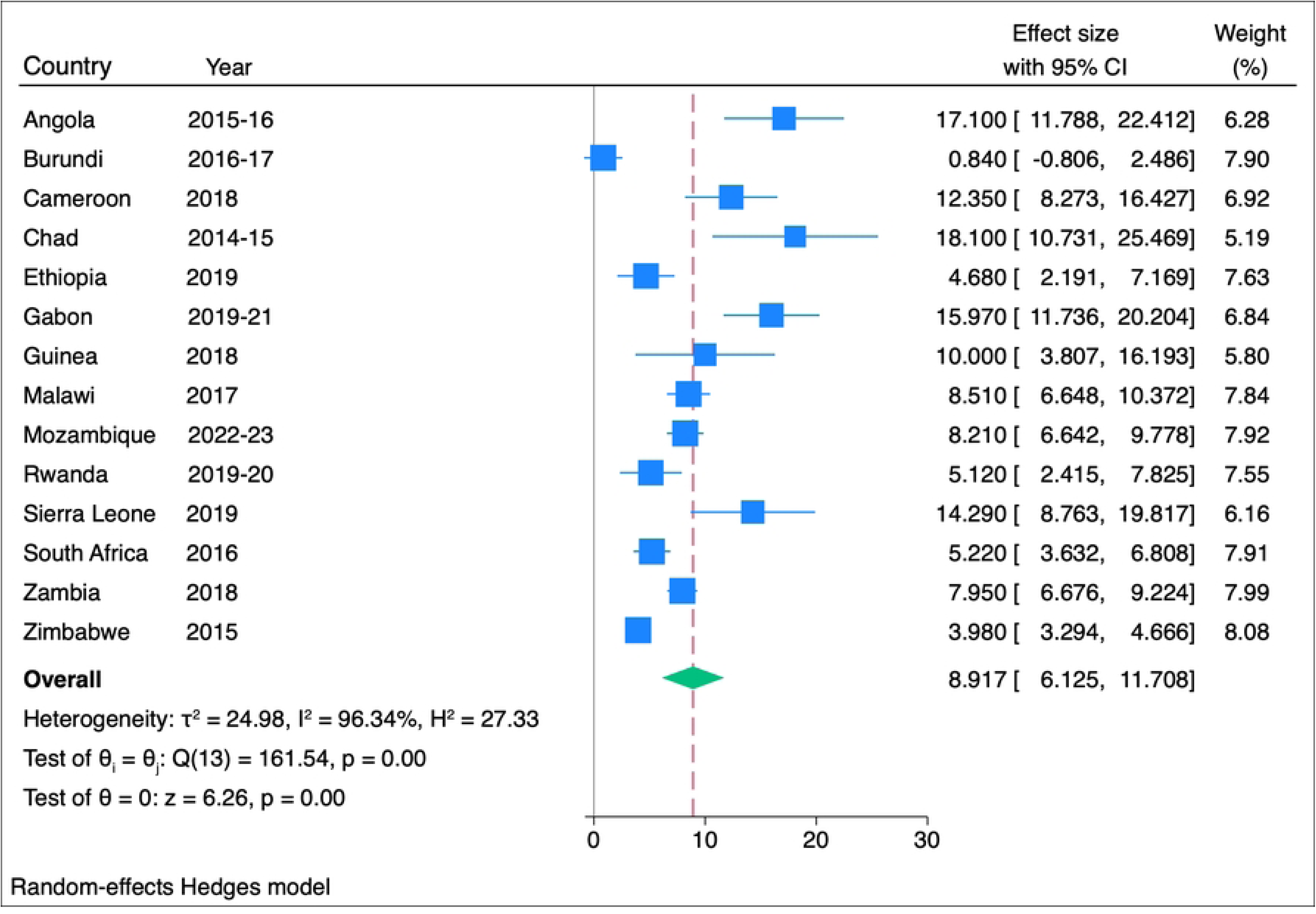
Unmet need for space.

**Figure 2.**
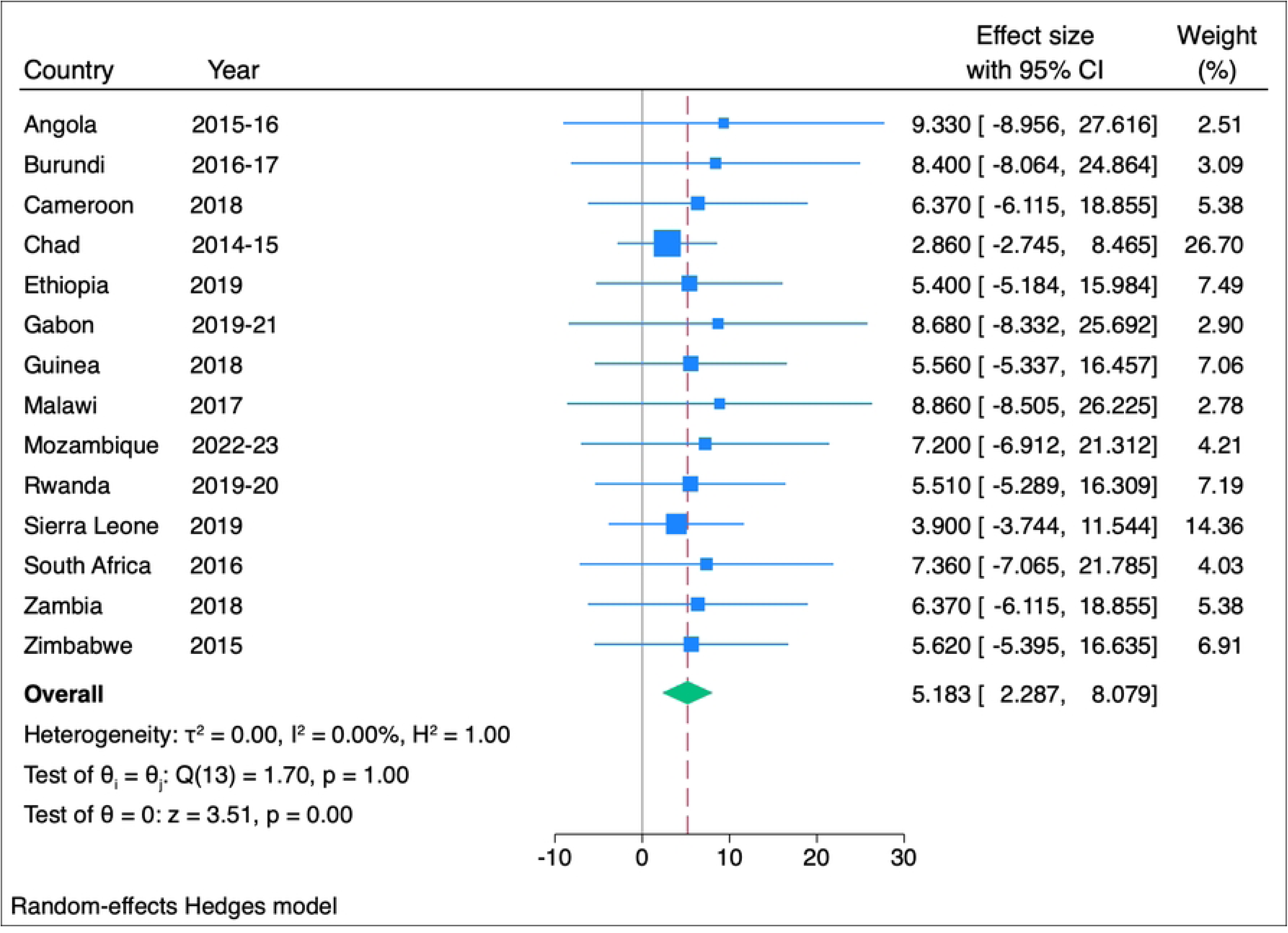
Unmet need for limiting.

**Figure 3.**
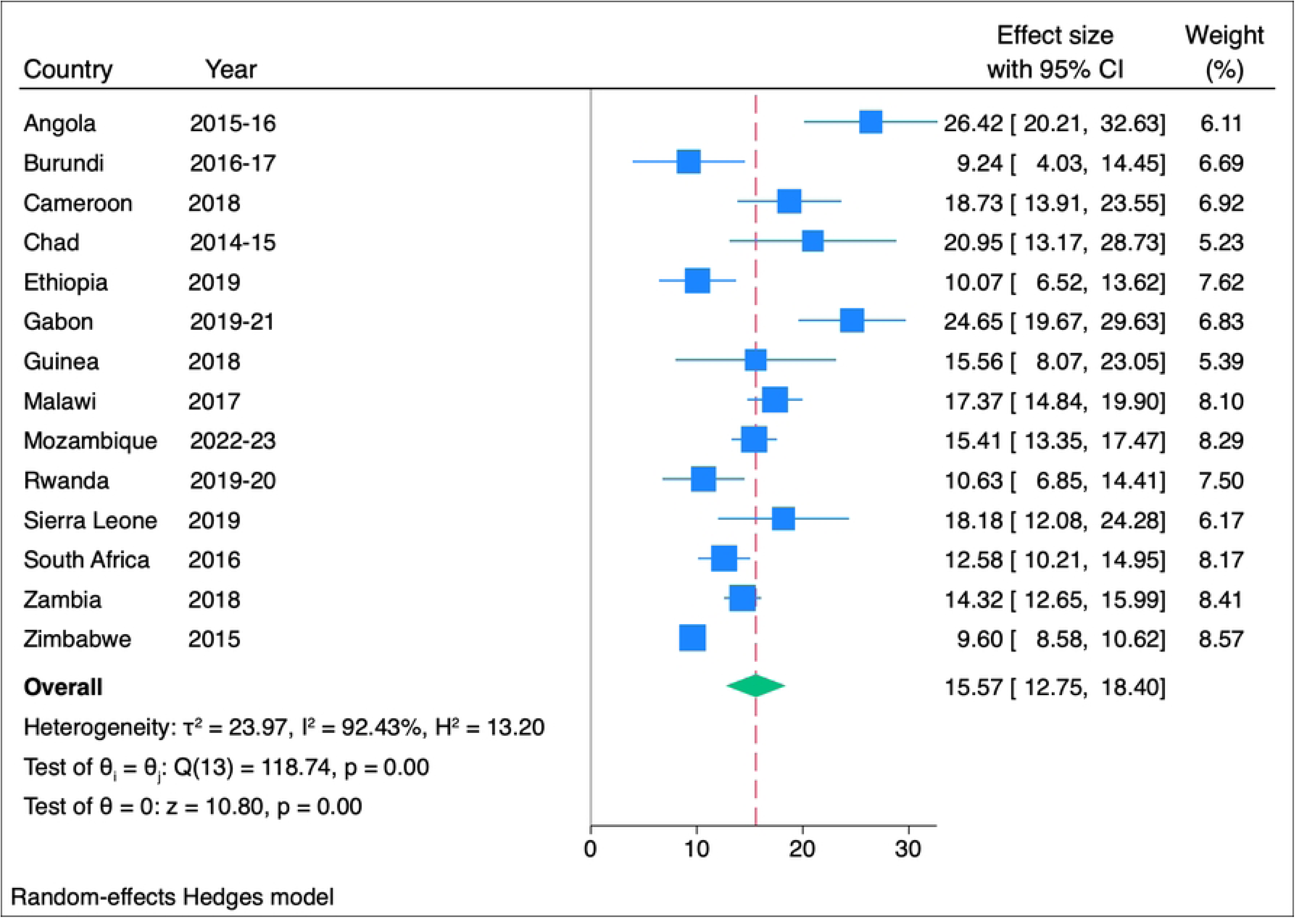
Unmet need for total family planning.

**Table 2:** Distribution of unmet needs of FP and their association with HIV status.

| Unmet need of Family Planning |  |  | HIV Status |  |
| --- | --- | --- | --- | --- |
|  | Frequency<br>(weighted) | Percent<br>(%) | Negative<br>(%) | Positive<br>(%) |
| **Unmet need for limiting** |  |  | **($\chi^2 = 37.895$, $p < 0.0001$)** |  |
| No | 100110 | 94.4 | 92.3 | 7.7 |
| Yes | 5960 | 5.6 | 90.2 | 9.8 |
| **Unmet need for spacing** |  |  | **($\chi^2 = 156.373$, $p < 0.0001$)** |  |
| No | 93,890 | 88.5 | 91.9 | 8.1 |
| Yes | 12,180 | 11.5 | 95.0 | 5.0 |
| **Unmet need total** |  |  | **($\chi^2 = 39.440$, $p < 0.0001$)** |  |
| No | 87,930 | 82.9 | 92.0 | 8.0 |
| Yes | 18,140 | 17.1 | 93.4 | 6.6 |

Table 3 shows the association between the unmet total needs of FP and women with positive HIV status. The study showed that there is significant association between all the socio-demographic characteristics and total unmet needs of family planning. Unmet need of FP was found to be associated with age, with 17.68% of women age between 20 – 24 years reporting unmet need of family planning. Among married women, 16.18% of them had unmet need of family planning. Also, 15.66% of women who ever used any contraceptive had total unmet needs of family planning. Also, 14.32% of the women living in rural area had total unmet need of family planning. It was again observed that 15.8% of women who indicated that distance to health facility was a big problem also had total unmet need of FP compared to 12.03% of women who said distance was not a big problem.

**Table 3:**
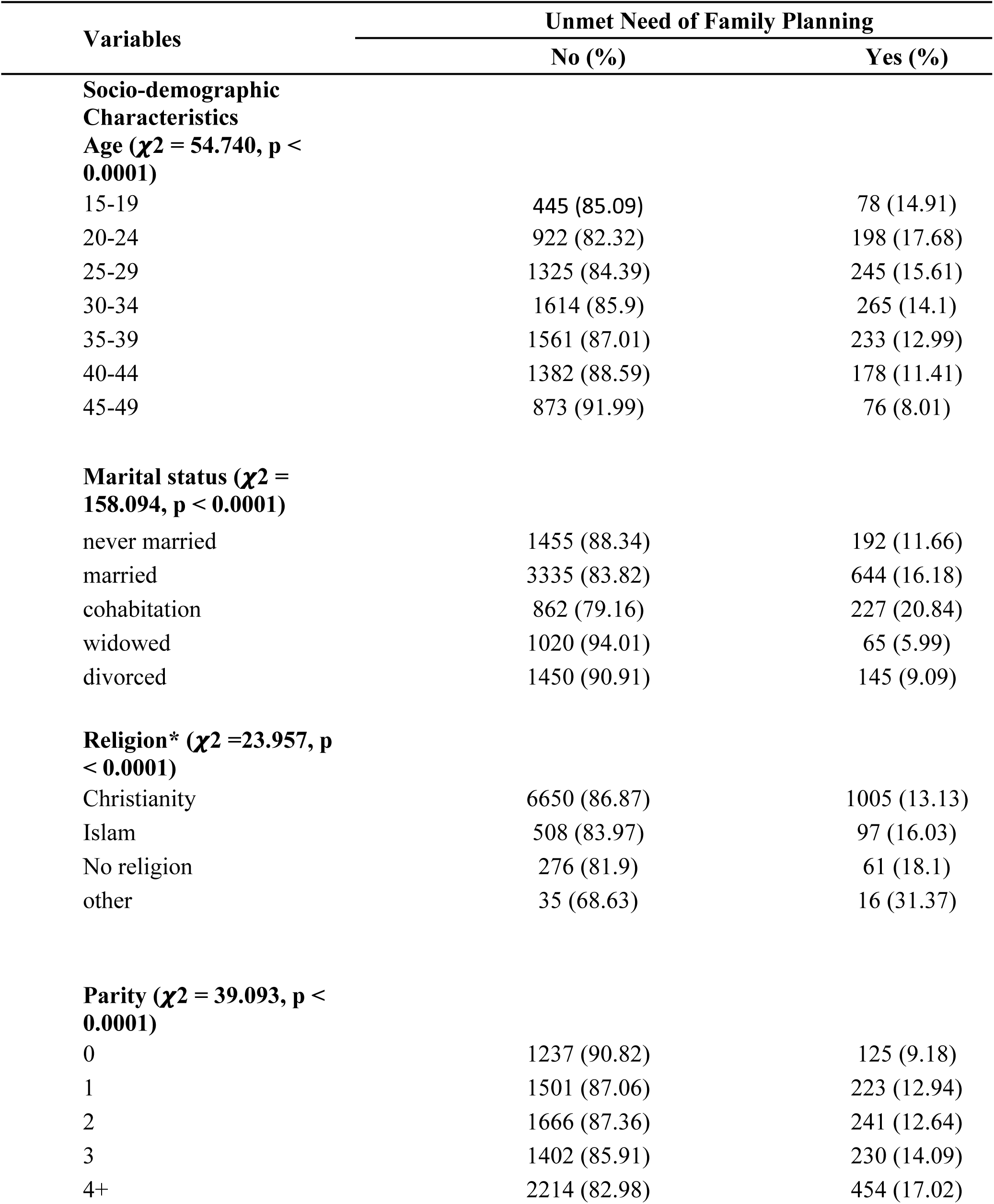
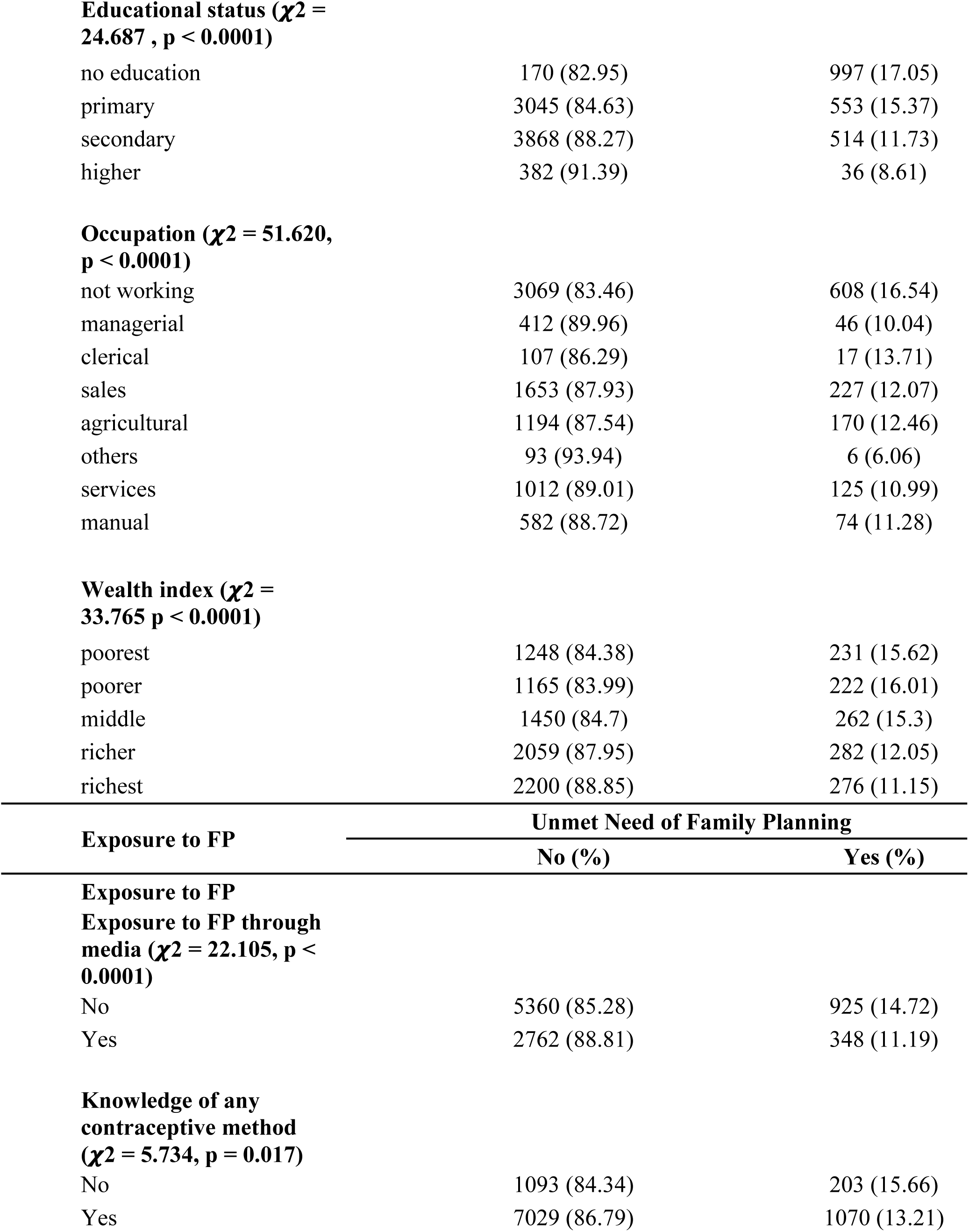
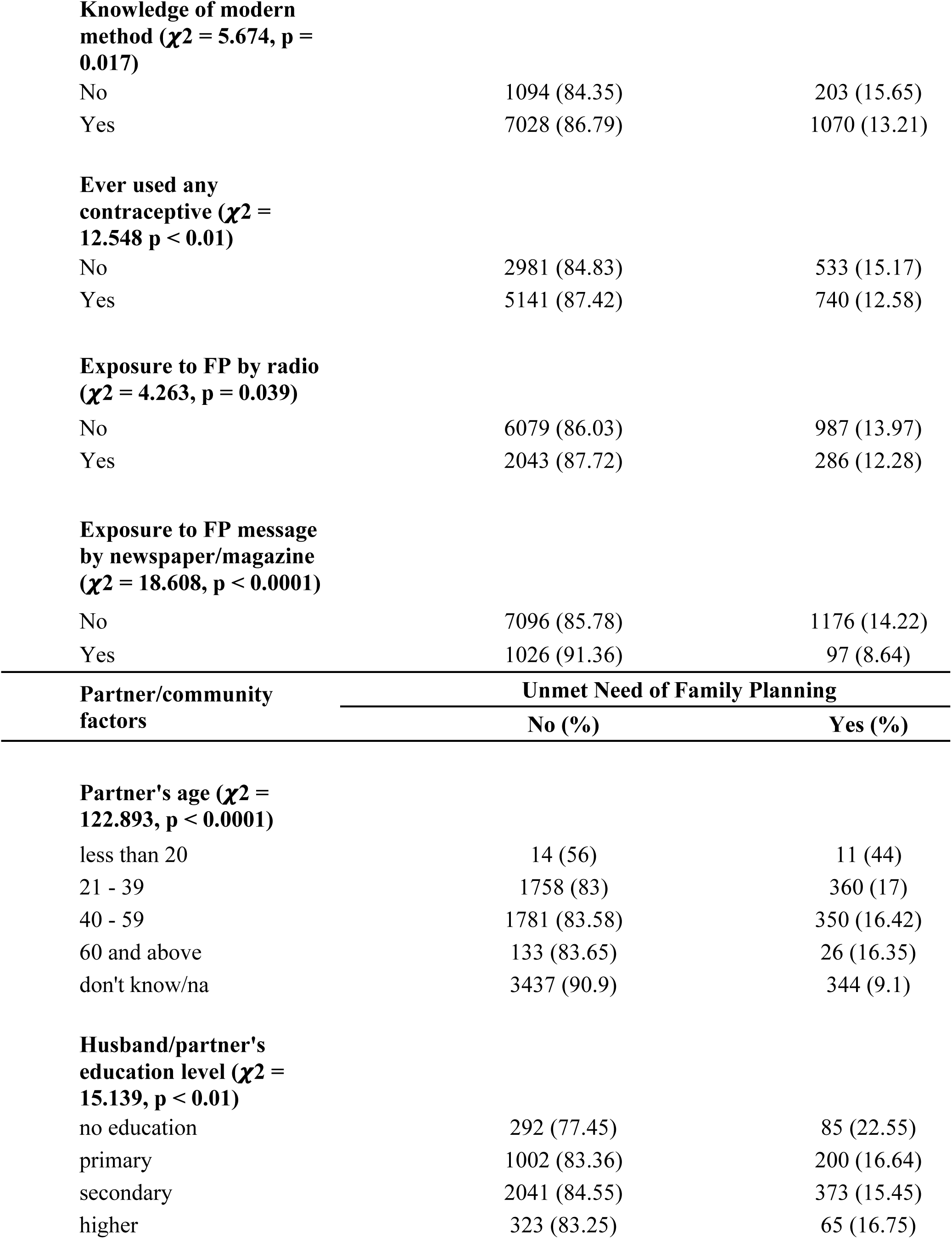
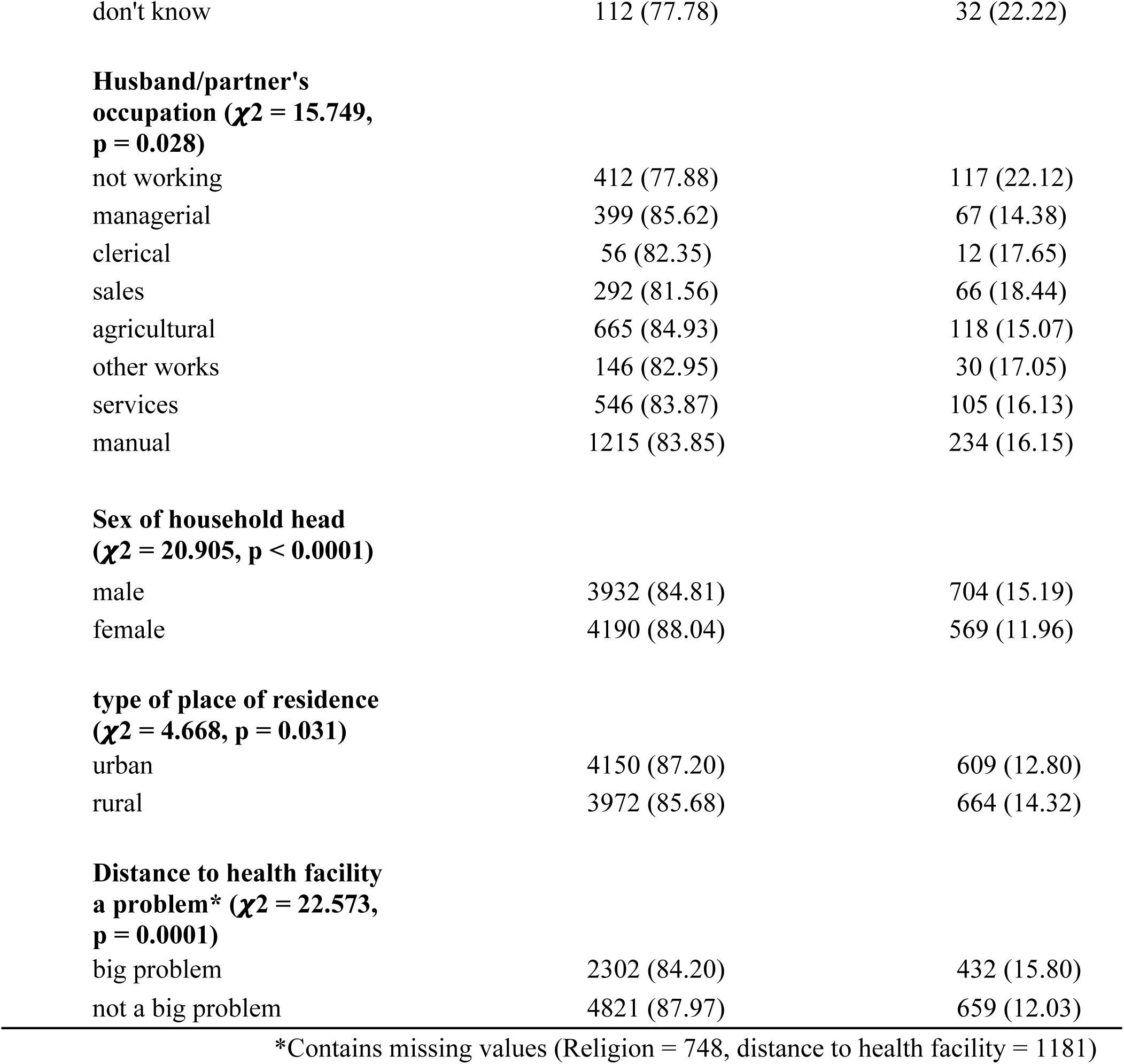
Explanatory variables and their association with unmet needs of FP among women living with HIV.

| Variables | Unmet Need of Family Planning |  |
| --- | --- | --- |
|  | No (%) | Yes (%) |
| **Socio-demographic Characteristics** |  |  |
| **Age ($\chi^2 = 54.740$, $p < 0.0001$)** |  |  |
| 15-19 | 445 (85.09) | 78 (14.91) |
| 20-24 | 922 (82.32) | 198 (17.68) |
| 25-29 | 1325 (84.39) | 245 (15.61) |
| 30-34 | 1614 (85.9) | 265 (14.1) |
| 35-39 | 1561 (87.01) | 233 (12.99) |
| 40-44 | 1382 (88.59) | 178 (11.41) |
| 45-49 | 873 (91.99) | 76 (8.01) |
| **Marital status ($\chi^2 = 158.094$, $p < 0.0001$)** |  |  |
| never married | 1455 (88.34) | 192 (11.66) |
| married | 3335 (83.82) | 644 (16.18) |
| cohabitation | 862 (79.16) | 227 (20.84) |
| widowed | 1020 (94.01) | 65 (5.99) |
| divorced | 1450 (90.91) | 145 (9.09) |
| **Religion\* ($\chi^2 = 23.957$, $p < 0.0001$)** |  |  |
| Christianity | 6650 (86.87) | 1005 (13.13) |
| Islam | 508 (83.97) | 97 (16.03) |
| No religion | 276 (81.9) | 61 (18.1) |
| other | 35 (68.63) | 16 (31.37) |
| **Parity ($\chi^2 = 39.093$, $p < 0.0001$)** |  |  |
| 0 | 1237 (90.82) | 125 (9.18) |
| 1 | 1501 (87.06) | 223 (12.94) |
| 2 | 1666 (87.36) | 241 (12.64) |
| 3 | 1402 (85.91) | 230 (14.09) |
| 4+ | 2214 (82.98) | 454 (17.02) |
| **Educational status ( $\chi^2 = 24.687$ , $p < 0.0001$ )** |  |  |
| no education | 170 (82.95) | 997 (17.05) |
| primary | 3045 (84.63) | 553 (15.37) |
| secondary | 3868 (88.27) | 514 (11.73) |
| higher | 382 (91.39) | 36 (8.61) |
| **Occupation ( $\chi^2 = 51.620$ , $p < 0.0001$ )** |  |  |
| not working | 3069 (83.46) | 608 (16.54) |
| managerial | 412 (89.96) | 46 (10.04) |
| clerical | 107 (86.29) | 17 (13.71) |
| sales | 1653 (87.93) | 227 (12.07) |
| agricultural | 1194 (87.54) | 170 (12.46) |
| others | 93 (93.94) | 6 (6.06) |
| services | 1012 (89.01) | 125 (10.99) |
| manual | 582 (88.72) | 74 (11.28) |
| **Wealth index ( $\chi^2 = 33.765$ , $p < 0.0001$ )** |  |  |
| poorest | 1248 (84.38) | 231 (15.62) |
| poorer | 1165 (83.99) | 222 (16.01) |
| middle | 1450 (84.7) | 262 (15.3) |
| richer | 2059 (87.95) | 282 (12.05) |
| richest | 2200 (88.85) | 276 (11.15) |

| Exposure to FP | Unmet Need of Family Planning |  |
| --- | --- | --- |
|  | No (%) | Yes (%) |
| **Exposure to FP** |  |  |
| **Exposure to FP through media ($\chi^2 = 22.105$, $p < 0.0001$)** |  |  |
| No | 5360 (85.28) | 925 (14.72) |
| Yes | 2762 (88.81) | 348 (11.19) |
| **Knowledge of any contraceptive method ($\chi^2 = 5.734$, $p = 0.017$)** |  |  |
| No | 1093 (84.34) | 203 (15.66) |
| Yes | 7029 (86.79) | 1070 (13.21) |
| **Knowledge of modern method ( $\chi^2 = 5.674$ , $p = 0.017$ )** |  |  |
| No | 1094 (84.35) | 203 (15.65) |
| Yes | 7028 (86.79) | 1070 (13.21) |
| **Ever used any contraceptive ( $\chi^2 = 12.548$ $p < 0.01$ )** |  |  |
| No | 2981 (84.83) | 533 (15.17) |
| Yes | 5141 (87.42) | 740 (12.58) |
| **Exposure to FP by radio ( $\chi^2 = 4.263$ , $p = 0.039$ )** |  |  |
| No | 6079 (86.03) | 987 (13.97) |
| Yes | 2043 (87.72) | 286 (12.28) |
| **Exposure to FP message by newspaper/magazine ( $\chi^2 = 18.608$ , $p < 0.0001$ )** |  |  |
| No | 7096 (85.78) | 1176 (14.22) |
| Yes | 1026 (91.36) | 97 (8.64) |

| Partner/community factors | Unmet Need of Family Planning |  |
| --- | --- | --- |
|  | No (%) | Yes (%) |
| **Partner's age ( $\chi^2 = 122.893$ , $p < 0.0001$ )** |  |  |
| less than 20 | 14 (56) | 11 (44) |
| 21 - 39 | 1758 (83) | 360 (17) |
| 40 - 59 | 1781 (83.58) | 350 (16.42) |
| 60 and above | 133 (83.65) | 26 (16.35) |
| don't know/na | 3437 (90.9) | 344 (9.1) |
| **Husband/partner's education level ( $\chi^2 = 15.139$ , $p < 0.01$ )** |  |  |
| no education | 292 (77.45) | 85 (22.55) |
| primary | 1002 (83.36) | 200 (16.64) |
| secondary | 2041 (84.55) | 373 (15.45) |
| higher | 323 (83.25) | 65 (16.75) |
| don't know | 112 (77.78) | 32 (22.22) |
| **Husband/partner's occupation ( $\chi^2 = 15.749$ , $p = 0.028$ )** |  |  |
| not working | 412 (77.88) | 117 (22.12) |
| managerial | 399 (85.62) | 67 (14.38) |
| clerical | 56 (82.35) | 12 (17.65) |
| sales | 292 (81.56) | 66 (18.44) |
| agricultural | 665 (84.93) | 118 (15.07) |
| other works | 146 (82.95) | 30 (17.05) |
| services | 546 (83.87) | 105 (16.13) |
| manual | 1215 (83.85) | 234 (16.15) |
| **Sex of household head ( $\chi^2 = 20.905$ , $p < 0.0001$ )** |  |  |
| male | 3932 (84.81) | 704 (15.19) |
| female | 4190 (88.04) | 569 (11.96) |
| **type of place of residence ( $\chi^2 = 4.668$ , $p = 0.031$ )** |  |  |
| urban | 4150 (87.20) | 609 (12.80) |
| rural | 3972 (85.68) | 664 (14.32) |
| **Distance to health facility a problem\* ( $\chi^2 = 22.573$ , $p = 0.0001$ )** |  |  |
| big problem | 2302 (84.20) | 432 (15.80) |
| not a big problem | 4821 (87.97) | 659 (12.03) |

\*Contains missing values (Religion = 748, distance to health facility = 1181)

Table 4 shows the effects of the individual, exposure to FP, and partner/community factors of women with HIV on unmet needs of FP. Among older age groups, the odds of having unmet needs decrease significantly; those aged 30-34, 35-39, and 40-44 have increasingly lower odds of unmet needs compared to the youngest age group across all models, with the strongest effect seen in the 45-49 age group [aOR = 0.159, CI = 0.0913-0.277 in Model 4]. Marital status also plays a critical role, with married individuals [aOR = 8.601, CI = 2.857-25.90] and those cohabiting [aOR = 9.096, CI = 3.017-27.42] showing significantly higher odds of unmet needs in Model 4, compared to those not married. Parity is strongly associated with unmet needs for family planning; as the number of children increases, so do the odds of having unmet needs. For instance, individuals with four or more children show the highest odds [aOR = 8.081, CI = 5.113-12.77 in Model 4], compared to women with no children. Exposure to FP through media [aOR = 0.564, CI = 0.468 - 0.678] was also found to significantly reduce odds of unmet needs of FP.

**Table 4:**
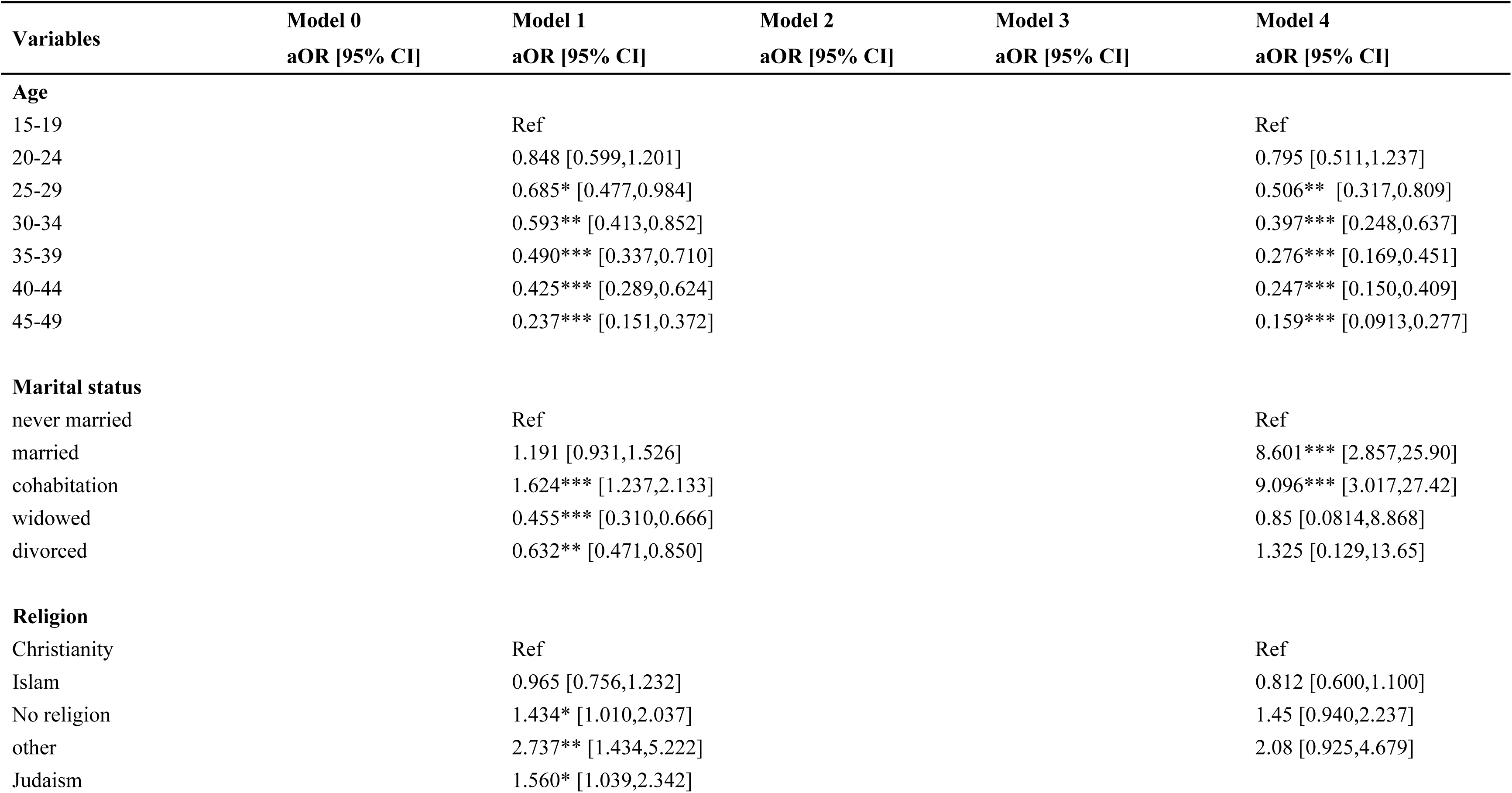
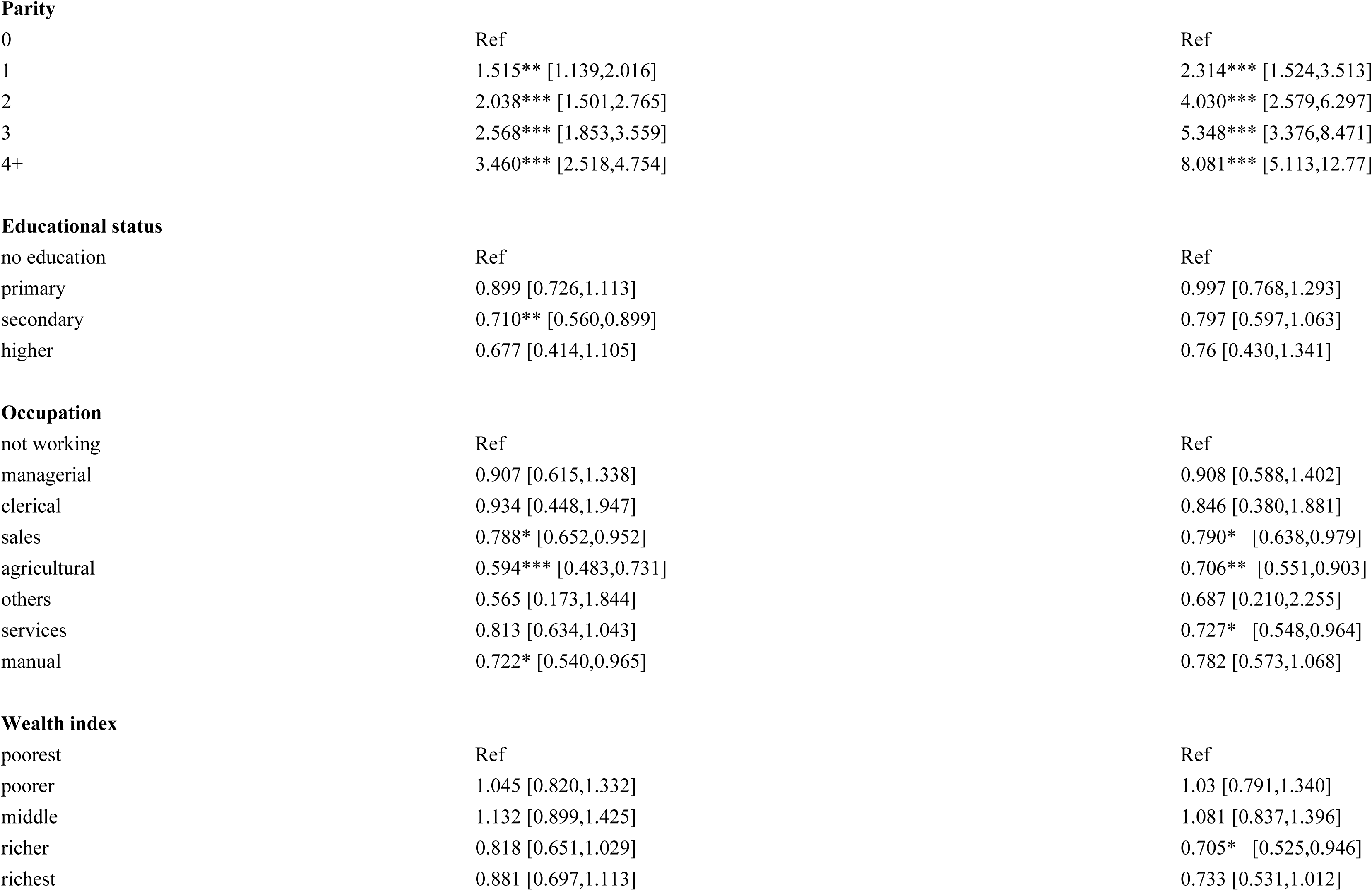
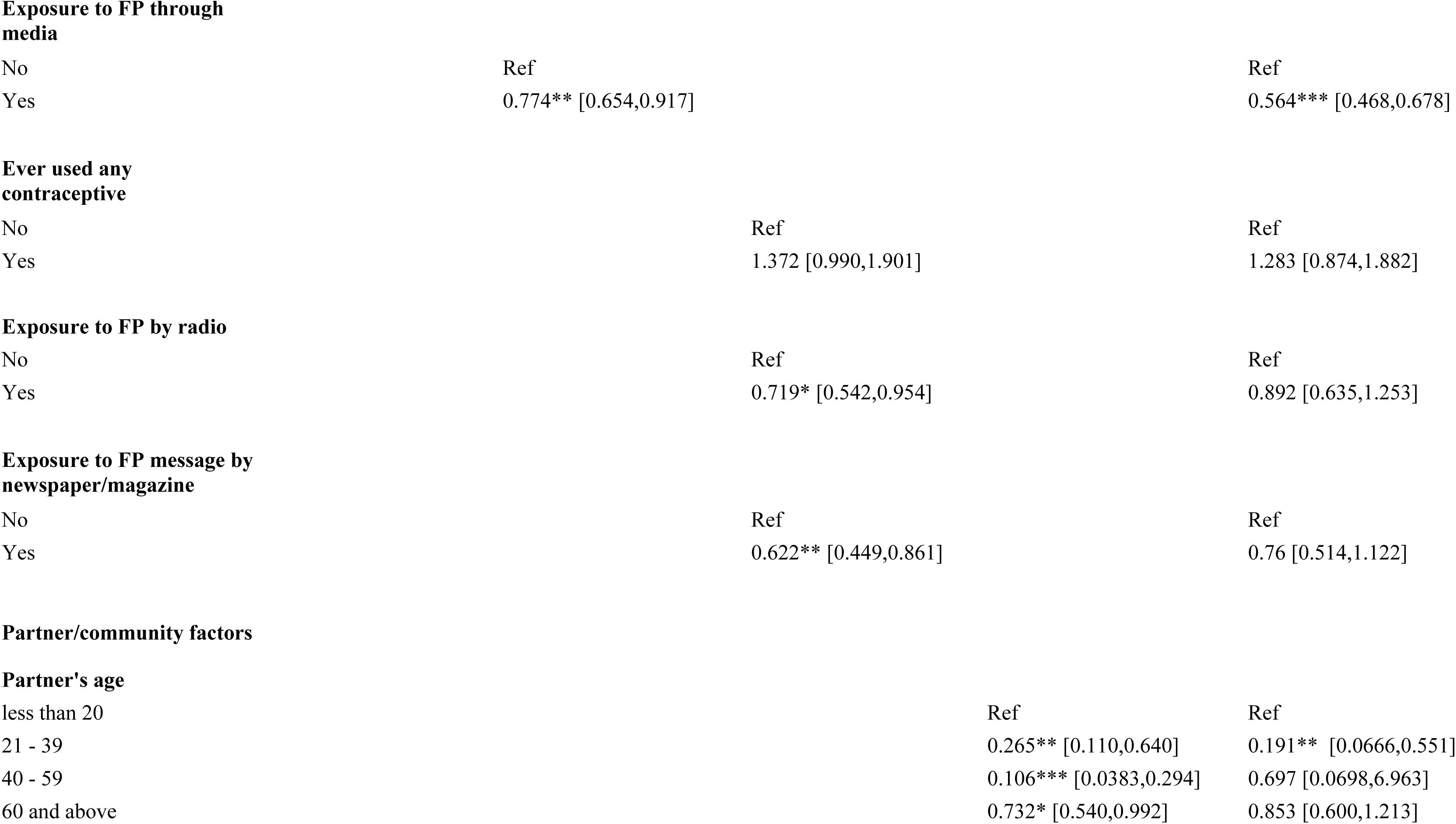
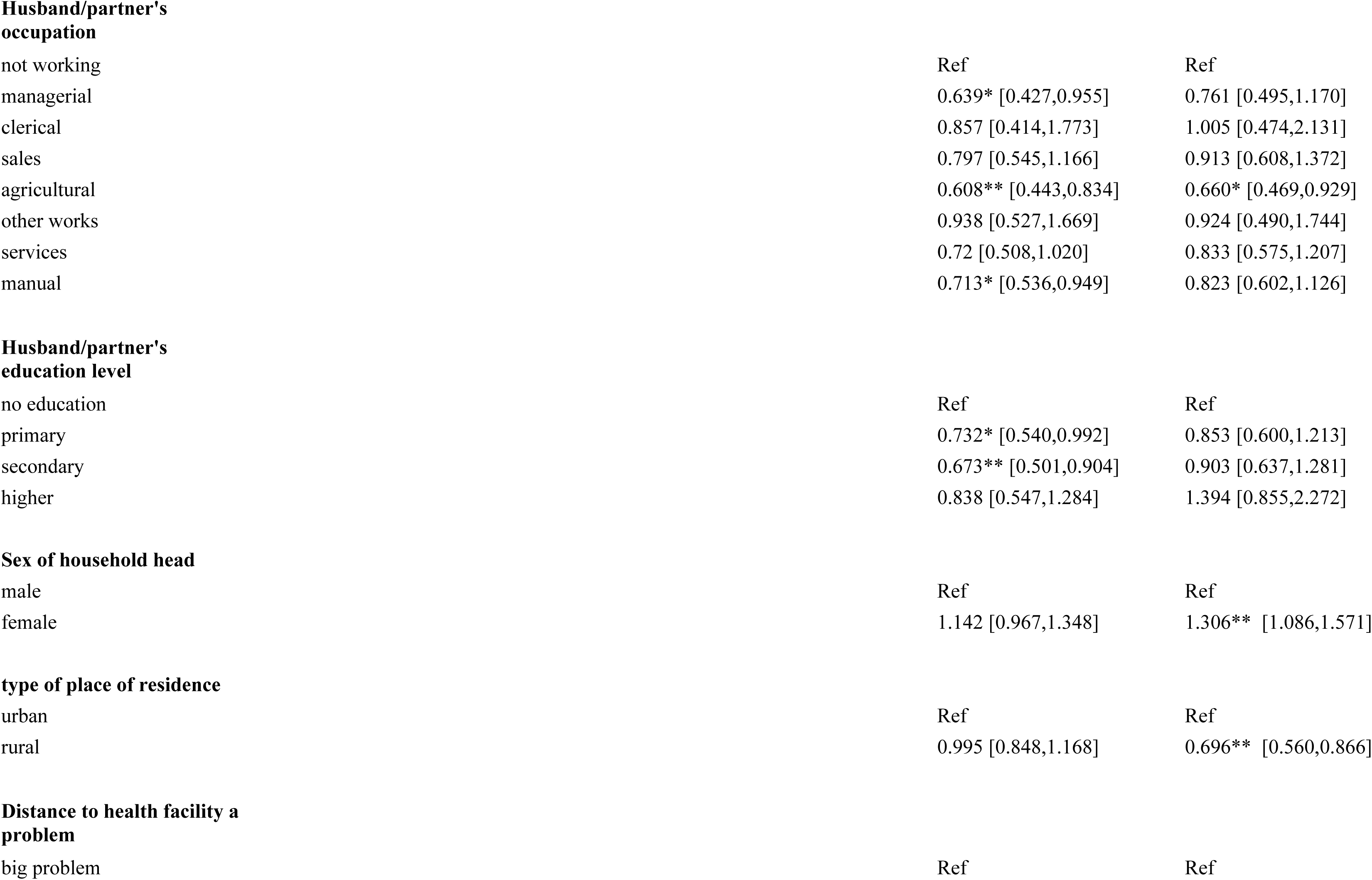
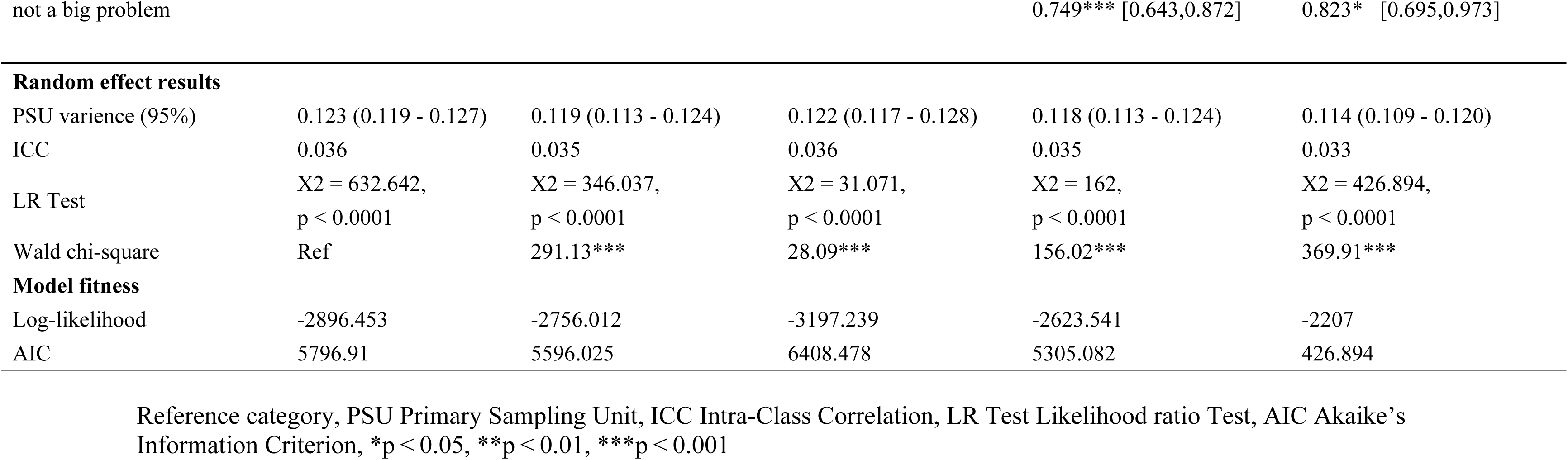
Mixed effects results of individual, exposure to FP, partner/community factors of women with HIV and their association with unmet need of family planning.

| Variables | Model 0<br>aOR [95% CI] | Model 1<br>aOR [95% CI] | Model 2<br>aOR [95% CI] | Model 3<br>aOR [95% CI] | Model 4<br>aOR [95% CI] |
| --- | --- | --- | --- | --- | --- |
| **Age** |  |  |  |  |  |
| 15-19 |  | Ref |  |  | Ref |
| 20-24 |  | 0.848 [0.599,1.201] |  |  | 0.795 [0.511,1.237] |
| 25-29 |  | 0.685* [0.477,0.984] |  |  | 0.506** [0.317,0.809] |
| 30-34 |  | 0.593** [0.413,0.852] |  |  | 0.397*** [0.248,0.637] |
| 35-39 |  | 0.490*** [0.337,0.710] |  |  | 0.276*** [0.169,0.451] |
| 40-44 |  | 0.425*** [0.289,0.624] |  |  | 0.247*** [0.150,0.409] |
| 45-49 |  | 0.237*** [0.151,0.372] |  |  | 0.159*** [0.0913,0.277] |
| **Marital status** |  |  |  |  |  |
| never married |  | Ref |  |  | Ref |
| married |  | 1.191 [0.931,1.526] |  |  | 8.601*** [2.857,25.90] |
| cohabitation |  | 1.624*** [1.237,2.133] |  |  | 9.096*** [3.017,27.42] |
| widowed |  | 0.455*** [0.310,0.666] |  |  | 0.85 [0.0814,8.868] |
| divorced |  | 0.632** [0.471,0.850] |  |  | 1.325 [0.129,13.65] |
| **Religion** |  |  |  |  |  |
| Christianity |  | Ref |  |  | Ref |
| Islam |  | 0.965 [0.756,1.232] |  |  | 0.812 [0.600,1.100] |
| No religion |  | 1.434* [1.010,2.037] |  |  | 1.45 [0.940,2.237] |
| other |  | 2.737** [1.434,5.222] |  |  | 2.08 [0.925,4.679] |
| Judaism |  | 1.560* [1.039,2.342] |  |  |  |

**Parity**
|  |  |  |
| --- | --- | --- |
| 0 | Ref | Ref |
| 1 | 1.515** [1.139,2.016] | 2.314*** [1.524,3.513] |
| 2 | 2.038*** [1.501,2.765] | 4.030*** [2.579,6.297] |
| 3 | 2.568*** [1.853,3.559] | 5.348*** [3.376,8.471] |
| 4+ | 3.460*** [2.518,4.754] | 8.081*** [5.113,12.77] |

**Educational status**
|  |  |  |
| --- | --- | --- |
| no education | Ref | Ref |
| primary | 0.899 [0.726,1.113] | 0.997 [0.768,1.293] |
| secondary | 0.710** [0.560,0.899] | 0.797 [0.597,1.063] |
| higher | 0.677 [0.414,1.105] | 0.76 [0.430,1.341] |

**Occupation**
|  |  |  |
| --- | --- | --- |
| not working | Ref | Ref |
| managerial | 0.907 [0.615,1.338] | 0.908 [0.588,1.402] |
| clerical | 0.934 [0.448,1.947] | 0.846 [0.380,1.881] |
| sales | 0.788* [0.652,0.952] | 0.790* [0.638,0.979] |
| agricultural | 0.594*** [0.483,0.731] | 0.706** [0.551,0.903] |
| others | 0.565 [0.173,1.844] | 0.687 [0.210,2.255] |
| services | 0.813 [0.634,1.043] | 0.727* [0.548,0.964] |
| manual | 0.722* [0.540,0.965] | 0.782 [0.573,1.068] |

**Wealth index**
|  |  |  |
| --- | --- | --- |
| poorest | Ref | Ref |
| poorer | 1.045 [0.820,1.332] | 1.03 [0.791,1.340] |
| middle | 1.132 [0.899,1.425] | 1.081 [0.837,1.396] |
| richer | 0.818 [0.651,1.029] | 0.705* [0.525,0.946] |
| richest | 0.881 [0.697,1.113] | 0.733 [0.531,1.012] |

**Exposure to FP through media**
|  |  |  |
| --- | --- | --- |
| No | Ref | Ref |
| Yes | 0.774** [0.654,0.917] | 0.564*** [0.468,0.678] |

**Ever used any contraceptive**
|  |  |  |
| --- | --- | --- |
| No | Ref | Ref |
| Yes | 1.372 [0.990,1.901] | 1.283 [0.874,1.882] |

**Exposure to FP by radio**
|  |  |  |
| --- | --- | --- |
| No | Ref | Ref |
| Yes | 0.719* [0.542,0.954] | 0.892 [0.635,1.253] |

**Exposure to FP message by newspaper/magazine**
|  |  |  |
| --- | --- | --- |
| No | Ref | Ref |
| Yes | 0.622** [0.449,0.861] | 0.76 [0.514,1.122] |

**Partner/community factors****Partner's age**
|  |  |  |
| --- | --- | --- |
| less than 20 | Ref | Ref |
| 21 - 39 | 0.265** [0.110,0.640] | 0.191** [0.0666,0.551] |
| 40 - 59 | 0.106*** [0.0383,0.294] | 0.697 [0.0698,6.963] |
| 60 and above | 0.732* [0.540,0.992] | 0.853 [0.600,1.213] |

**Husband/partner's occupation**
|  |  |  |
| --- | --- | --- |
| not working | Ref | Ref |
| managerial | 0.639* [0.427,0.955] | 0.761 [0.495,1.170] |
| clerical | 0.857 [0.414,1.773] | 1.005 [0.474,2.131] |
| sales | 0.797 [0.545,1.166] | 0.913 [0.608,1.372] |
| agricultural | 0.608** [0.443,0.834] | 0.660* [0.469,0.929] |
| other works | 0.938 [0.527,1.669] | 0.924 [0.490,1.744] |
| services | 0.72 [0.508,1.020] | 0.833 [0.575,1.207] |
| manual | 0.713* [0.536,0.949] | 0.823 [0.602,1.126] |

**Husband/partner's education level**
|  |  |  |
| --- | --- | --- |
| no education | Ref | Ref |
| primary | 0.732* [0.540,0.992] | 0.853 [0.600,1.213] |
| secondary | 0.673** [0.501,0.904] | 0.903 [0.637,1.281] |
| higher | 0.838 [0.547,1.284] | 1.394 [0.855,2.272] |

**Sex of household head**
|  |  |  |
| --- | --- | --- |
| male | Ref | Ref |
| female | 1.142 [0.967,1.348] | 1.306** [1.086,1.571] |

**type of place of residence**
|  |  |  |
| --- | --- | --- |
| urban | Ref | Ref |
| rural | 0.995 [0.848,1.168] | 0.696** [0.560,0.866] |

**Distance to health facility a problem**
|  |  |  |
| --- | --- | --- |
| big problem | Ref | Ref |
| **Random effect results** |  |  |  |  |  |
| PSU variance (95%) | 0.123 (0.119 - 0.127) | 0.119 (0.113 - 0.124) | 0.122 (0.117 - 0.128) | 0.118 (0.113 - 0.124) | 0.114 (0.109 - 0.120) |
| ICC | 0.036 | 0.035 | 0.036 | 0.035 | 0.033 |
| LR Test | X2 = 632.642,<br>p < 0.0001 | X2 = 346.037,<br>p < 0.0001 | X2 = 31.071,<br>p < 0.0001 | X2 = 162,<br>p < 0.0001 | X2 = 426.894,<br>p < 0.0001 |
| Wald chi-square | Ref | 291.13*** | 28.09*** | 156.02*** | 369.91*** |
| **Model fitness** |  |  |  |  |  |
| Log-likelihood | -2896.453 | -2756.012 | -3197.239 | -2623.541 | -2207 |
| AIC | 5796.91 | 5596.025 | 6408.478 | 5305.082 | 426.894 |

Reference category, PSU Primary Sampling Unit, ICC Intra-Class Correlation, LR Test Likelihood ratio Test, AIC Akaike's Information Criterion, \*p < 0.05, \*\*p < 0.01, \*\*\*p < 0.001

Community factors such as sex of household head, type of place of residence, and distance to health facility significantly affected the unmet needs for family planning. Household heads who were females showed a higher odd of unmet needs of FP compared to male household heads [aOR = 1.306, CI = 1.086 - 1.571]. Women living in rural area showed a lower odds of unmet need of FP, compared to those living in urban areas [aOR = 0.696, CI = 0.560 - 0.866].

## DISCUSSION

The concept of unmet need for contraception is crucial for developing effective FP programs, as it has significant implications for the maternal and child health, particularly among women of reproductive age living with HIV. This study assessed the extents of unmet need for FP among women of reproductive age living with HIV in SSA taking into consideration the various factors like the individual factors, exposure to FP services, and partner/community factors that could influence the uptake of FP among this vulnerable group. The combination of these multilevel factors is critical for a holistic assessment of unmet needs of FP among these women in SSA.

This study found that 15.57% of the WRLWH included in the study had an total unmet need for family planning. This finding somehow concurs with a study conducted in the Oromia regional state of Ethiopia identified that 16% of women of reproductive age living with HIV had an unmet need for family planning, highlighting the demand for integrated HIV and FP services (24). Other study conducted in the South-South region of Nigeria found a higher prevalence of unmet FP needs among people living with HIV/AIDS, with 43.5% of the study population experiencing unmet needs (25). The high rate of unmet need of FP among this active reproductive age group of women increases the risk of mother-to-child transmission of HIV during pregnancy, childbirth, or breastfeeding if not properly managed with antiretroviral therapy.

The analysis of factors of unmet need FP among HIV-positive women in our study indicated that age of the individuals involved in the study emerged as a significant factor to the unmet demand for FP services among women living with HIV in SSA. Among older age groups, the odds of having unmet needs decrease significantly. Those aged 30-34, 35-39, and 40-44 have increasingly lower odds of unmet needs compared to the youngest age group across all models. As women age, their fertility goals often change. Older women may desire fewer children or no additional children, which can lead to a more deliberate and effective use of contraception. This shift in reproductive intentions could reduce the incidence of unmet needs. This finding is in contrast with a study done in Nigeria which found that age group was significantly associated with unmet FP needs, with those aged 40 years or older having higher odds of unmet needs (26). A study in Ethiopia identified that women below 18 years at first marriage had increased odds of unmet FP needs, supporting our current study (25). Additionally, a study in Zambia highlighted age of women as a predictor of unmet need for FP among women living with HIV, emphasizing the importance of considering age in addressing this issue (27). This study findings suggest a tailored FP programs according to age-specific needs and challenges. Younger women may need more education and outreach to understand and access FP options.

The study found that married individuals and those cohabiting have significantly higher odds of unmet needs among WLWH in SSA, compared to those not in any union. Married or cohabiting women are likely to be more sexually active compared to their single counterparts. This increased frequency of sexual activity may lead to a higher demand for FP methods, which, if not met, increases the reported unmet need. Also, women in unions may face challenges in negotiating contraceptive use with their partners. If their partners oppose or are not supportive of contraceptive use, this can lead to a higher unmet need. Additionally, issues of power dynamics within the relationship can affect a woman’s autonomy in making health-related decisions.

Also, our study findings suggest that higher parity has the higher chances of unmet needs for family planning. For instance, individuals with four or more children show the highest odds, compared to those with no children. This finding corresponds with a study conducted in Ethiopia found that women with four or more children had 3.25 times higher odds of having unmet needs for FP compared to women with no children (28). Other study in Kenya reported that women with four or more children had 2.6 times higher odds of having unmet needs for FP compared to women with no children (29). Another study in India found that women with four or more children had 4.2 times higher odds of having unmet needs for FP compared to women with no children (30). The findings from these studies corroborate the strong association between higher parity and increased unmet needs for family planning, although the magnitude of the association may vary across different contexts and populations. This study finding could be attributed that as the number of children increases, the need for effective contraception to prevent further pregnancies also increases. Women with many children may have reached or exceeded their desired family size and thus actively seek to avoid additional pregnancies.

Regarding exposure to FP services and information, our study finding show that individuals who were exposed to FP information through media had 43.6% lower odds of having unmet needs for FP among WLWH in SSA. The result suggests that media-based interventions and campaigns for FP can be an effective strategy to address unmet needs. Through disseminating information about FP options, benefits, and access through various media channels (e.g., television, radio, social media, etc.), individuals may be better informed and more empowered to meet their FP needs. The study finding aligns with similar research in Sierra Leone, where lack of media exposure was significantly associated with a higher prevalence of unmet need for FP among adolescent girls and young women (31).

Understanding the partner and community dynamic are important to understanding dynamics of unmet needs of FP in SSA. In SSA, partners, especially male partners, are the gate keepers and decision makers for their female partners. Their decisions largely influence access to healthcare services among women. Also, the socio-culture and economic characteristics of communities in SSA have influence in the availability and access to maternal healthcare services. Our study looked into these partner and community factors that could influence unmet needs of FP among WLWH in SSA. Interestingly, our study found that female-headed households showed a higher likelihood of unmet FP needs compared to male-headed households. Female-headed households are often economically disadvantaged compared to male-headed households. This economic disparity can affect access to healthcare services, including family planning. Limited financial resources mean that these women may not be able to afford transportation to clinics, the cost of contraceptives, or even the associated medical consultations.

Also, this study found women living with HIV/AIDS in rural areas have lower odds of having unmet needs for family planning, compared to their counterparts in urban areas, suggesting that rural women living with HIV/AIDS may have better access to or higher utilization of FP services, despite the general challenges associated with accessing healthcare in rural settings. The result shows a potential disparity in FP access between rural and urban settings. A study in Malawi found that rural women living with HIV/AIDS had higher odds of using modern contraceptives compared to their urban counterparts (32). These are result of targeted efforts by public health programs to improve access and utilization of FP services in rural areas, especially for vulnerable populations like those living with HIV/AIDS.

## CONCLUSION

This study highlights the significant factors influencing unmet needs for FP among WRLWH in SSA. The findings reveal that individual factors such as age, marital status, and parity, as well as exposure to family planning information through media, play crucial roles in determining unmet needs. Additionally, partner and community dynamics, including household headship and rural versus urban residence, also impact access to and utilization of family planning services.

The study emphasizes the importance of tailored FP programs that consider the diverse needs and challenges faced by women living with HIV. Targeted interventions, particularly those focusing on media-based education and outreach, could effectively reduce unmet needs for FP among this vulnerable population. Addressing these unmet needs is essential for improving maternal and child health outcomes, PMTCT of HIV, and achieving the SDG related to health and gender equality in the region.

By identifying the predictors of unmet FP needs, this research provides valuable insights for developing comprehensive and effective public health strategies. These strategies should empower women living with HIV to make informed decisions about their reproductive health, ultimately enhancing their well-being and contributing to better health outcomes for their families and communities.

## Declaration

### Funding

No funding was received for conducting this study.

## Data Availability

Data for this study is freely available on the DHS program website (https://dhsprogram.com/Data/)

## Acknowledgements

We would like to thank the Measure DHS Program for providing the DHS datasets.

## Author Contributions

**Conceptualization** - MHK, & MMM; Methodology – MHK, & SSO; Data Curation: MHK, & DP; Formal analysis – MHK, TC & SSO; Writing of Original draft – MMM & ELM; Writing review & editing – MMM, ELM, & DP; Supervision & Validation – AAA.

## Notes

### Competing Interest Statement

The authors have declared no competing interest.

### Funding Statement

This study did not receive any form of funding

## REFERENCES

1. Kumah E, Boakye DS, Boateng R, Agyei E. Advancing the Global Fight Against HIV/Aids: Strategies, Barriers, and the Road to Eradication. Ann Glob Heal. United States; 2023;89(1):83.

2. World Health Organization. HIV and AIDS [Internet]. Website. 2023 [cited 2024 Jun 11]. Available from: https://www.who.int/news-room/fact-sheets/detail/hiv-aids?

3. World Health Organization. THE GLOBAL HEALTH OBSERVATORY [Internet]. Website. 2024 [cited 2024 Jun 10]. Available from: https://www.who.int/data/gho/data/indicators/indicator-details/GHO

4. Nkhoma L, Sitali DC, Zulu JM. Integration of family planning into HIV services: a systematic review. Ann Med [Internet]. Taylor & Francis; 2022;54(1):393–403. Available from: 10.1080/07853890.2021.2020893

5. Wilcher R, Petruney T, Cates W. The role of family planning in elimination of new pediatric HIV infection. Curr Opin HIV AIDS. 2013;8(5):490–7.

6. Chola L, McGee S, Tugendhaft A, Buchmann E, Hofman K. Scaling up family planning to reduce maternal and child mortality: The potential costs and benefits of modern contraceptive use in South Africa. PLoS One. 2015;10(6):1–16.

7. Wai MM, Bjertness E, Stigum H, Htay TT, Liabsuetrakul T, Myint ANM, et al. Unmet need for family planning among urban and rural married women in yangon region, myanmar—a cross-sectional study. Int J Environ Res Public Health. 2019;16(19).

8. Agyekum AK, Adde KS, Aboagye RG, Salihu T, Seidu AA, Ahinkorah BO. Unmet need for contraception and its associated factors among women in Papua New Guinea: analysis from the demographic and health survey. Reprod Health [Internet]. BioMed Central; 2022;19(1):1–11. Available from: 10.1186/s12978-022-01417-7

9. Pillai VK, Nagoshi JL. Unmet Family Planning Need Globally: A Clarion Call for Sharpening Current Research Frame Works. Open Access J Contracept. 2023;Volume 14(July):139–47.

10. Mavodza C V., Busza J, Mackworth-Young CRS, Nyamwanza R, Nzombe P, Dauya E, et al. Family Planning Experiences and Needs of Young Women Living With and Without HIV Accessing an Integrated HIV and SRH Intervention in Zimbabwe-An Exploratory Qualitative Study. Front Glob Women’s Heal. 2022;3(May):1–11.

11. Haberlen SA, Narasimhan M, Beres LK, Kennedy CE. Integration of Family Planning Services into HIV Care and Treatment Services: A Systematic Review. Stud Fam Plann. 2017;48(2):153–77.

12. Tomilola OR, Bolanle OE, Olufunmilayo OM. Unintended pregnancies among HIV-positive women in sub-Saharan Africa: a scoping review protocol. Syst Rev [Internet]. BioMed Central; 2023;12(1):1–6. Available from: 10.1186/s13643-023-02168-7

13. Lungu EA, Chewe M. Trends and predictors of unmet need for family planning among women living with HIV in Zambia: implications for elimination of vertical transmission of HIV. BMC Public Health. 2024;24(1):1–14.

14. Hagey JM, Akama E, Ayieko J, Bukusi EA, Cohen CR, Patel RC. Barriers and facilitators adolescent females living with HIV face in accessing contraceptive services: A qualitative assessment of providers’ perceptions in western Kenya. J Int AIDS Soc. 2015;18(1):1–8.

15. Elmi N, Marquez NG, Rucinski K, Lyons C, Turpin G, Ba I, et al. Meeting the reproductive health needs of female sex workers in Côte d’Ivoire: protecting the human right to dignified health. Reprod Health. 2023;20(1):1–10.

16. Behaviors SR, Ellerson RM. Health Services Utilization Among South African. AIDS Patient Care STDS. 2010;24(4).

17. Prata N. Making family planning accessible in resource-poor settings. Philos Trans R Soc B Biol Sci. 2009;364(1532):3093–9.

18. Dockalova B, Lau K, Barclay H, Marshall A. Sustainable development goals and family planning 2020. The International Planned Parenthood Federation (IPPF) United Kingdom. 2016:1–12.

19. DHS Program, USAID. The Demographic and Health Survey Program.

20. Corsi DJ, Neuman M, Finlay JE, Subramanian S V. Demographic and health surveys: A profile. Int J Epidemiol. 2012 Dec;41(6):1602–13.

21. Cleland J, Harbison S, Shah IH. Unmet Need for Contraception: Issues and Challenges. Stud Fam Plann [Internet]. 2014 Jun 1;45(2):105–22. Available from: 10.1111/j.1728-4465.2014.00380.x

22. Rossier C, Bradley SEK, Ross J, Winfrey W. Reassessing Unmet Need for Family Planning in the Postpartum Period. Stud Fam Plann. 2015;46(4):355–67.

23. Clarke P, Crawford C, Steele F, Vignoles A. Revisiting fixed-and random-effects models: some considerations for policy-relevant education research. Educ Econ. 2015;23(3):259– 77.

24. Ammie M, Arefaynie M, Adane B, Hussein K, Hassan M. Determinants of unmet need for family planning among currently married reproductive age women at Dewa Chefa District of Oromia special zone, Amhara region, Ethiopia, 2021; a case-control study. BMC Womens Health. 2024;24(1).

25. Assefa AA, Selassie SG, Mesele A, Kebede HB, Fikrie A, Abera G. Unmet need for family planning and associated factors among currently married women in Hawella Tulla subcity, Hawassa, southern Ethiopia: community-based study. Contracept Reprod Med [Internet]. 2023;8(1):1–10. Available from: 10.1186/s40834-022-00212-w

26. Ofurum IC, Mba OG, Enyindah CE. Factors Associated with Unmet Needs for Family Planning among People Living with HIV/AIDS in the South-South Region of Nigeria. J Adv Med Pharm Sci. 2023;25(1):10–22.

27. Lungu EA, Chewe M. Trends and predictors of unmet need for family planning among women living with HIV in Zambia: implications for elimination of vertical transmission of HIV. BMC Public Health. 2024;24(1).

28. Gebre G, Birhan N, Gebreslasie K. Prevalence and factors associated with unmet need for family planning among the currently married reproductive age women in shire-Enda- Slassie, northern west of Tigray, Ethiopia 2015: A community based cross-sectional study. Pan Afr Med J. 2016;23:1–9.

29. Ochako R, Temmerman M, Mbondo M, Askew I. Determinants of modern contraceptive use among sexually active men in Kenya. Reprod Health. 2017;14(1):1–15.

30. Devaraj K, Gausman J, Mishra R, Kumar A, Kim R, Subramanian S V. Trends in prevalence of unmet need for family planning in India: patterns of change across 36 States and Union Territories, 1993–2021. Reprod Health [Internet]. 2024;21(1):1–13. Available from: 10.1186/s12978-024-01781-6

31. Sserwanja Q, Turimumahoro P, Nuwabaine L, Kamara K, Musaba MW. Association between exposure to family planning messages on different mass media channels and the utilization of modern contraceptives among young women in Sierra Leone: insights from the 2019 Sierra Leone Demographic Health Survey. BMC Womens Health [Internet]. 2022;22(1):1–10. Available from: 10.1186/s12905-022-01974-w

32. Habte D, Namasasu J. Family planning use among women living with HIV: Knowing HIV positive status helps - Results from a national survey. Reprod Health. 2015;12(1):1– 11

